# A Systematic Approach to Identify Neuroprotective Interventions for Motor Neuron Disease

**DOI:** 10.1101/2022.04.13.22273823

**Authors:** The Writing Committee for Repurposing Living Systematic Review – Motor Neuron Disease (ReLiSyR-MND), Charis Wong, Jenna M. Gregory, Jing Liao, Kieren Egan, Hanna M. Vesterinen, Aimal Ahmad Khan, Maarij Anwar, Caitlin Beagan, Fraser Brown, John Cafferkey, Alessandra Cardinali, Jane Yi Chiam, Claire Chiang, Victoria Collins, Joyce Dormido, Elizabeth Elliott, Peter Foley, Yu Cheng Foo, Lily Fulton-Humble, Angus B. Gane, Stella A. Glasmacher, Áine Heffernan, Kiran Jayaprakash, Nimesh Jayasuriya, Amina Kaddouri, Jamie Kiernan, Gavin Langlands, Danielle Leighton, Jiaming Liu, James Lyon, Arpan R. Mehta, Alyssa Meng, Vivienne Nguyen, Na Hyun Park, Suzanne Quigley, Yousuf Rashid, Andrea Salzinger, Bethany Shiell, Ankur Singh, Tim Soane, Alexandra Thompson, Olaf Tomala, Fergal M. Waldron, Bhuvaneish T. Selvaraj, Jeremy Chataway, Robert Swingler, Peter Connick, Suvankar Pal, Siddharthan Chandran, Malcolm R. Macleod

## Abstract

**Background:** Motor neuron disease (MND) is an incurable progressive neurodegenerative disease with limited treatment options. There is a pressing need for innovation in identifying therapies to take to clinical trial.

**Objectives:** Here we detail a systematic, structured, and unbiased evidence-based approach to guide selection of drugs for clinical evaluation in the Motor Neuron Disease – Systematic Multi-arm Adaptive Randomised Trial (MND-SMART, clinicaltrials.gov registration number: NCT04302870), an adaptive platform trial.

**Methods:** We conducted a two-stage systematic review and meta-analysis to identify potential neuroprotective interventions. In stage one, we identified drugs from the clinical literature tested in at least one study in MND or in two or more cognate diseases with potential shared pivotal pathways (Alzheimer’s disease, Huntington’s disease, Parkinson’s disease, or multiple sclerosis). We scored and ranked 66 drugs thus identified using a predefined framework evaluating safety, efficacy, study size and quality of studies. In stage two, we conducted a systematic review of the MND preclinical literature describing efficacy of these drugs in animal models, multicellular eukaryotic models and human induced pluripotent stem cell studies; 17 of these drugs were reported to improve survival in at least one preclinical study. An expert panel then shortlisted and ranked 22 drugs considering stage one and stage two findings, mechanistic plausibility, safety and tolerability, findings from previous clinical trials in MND, and feasibility for use in clinical trials.

**Results:** Based on this process, the panel selected memantine and trazodone for testing in MND-SMART.

**Discussion:** For future drug selection, we will incorporate automation tools, text-mining and machine learning techniques to the systematic reviews and consider data generated from other domains, including high-throughput phenotypic screening of human induced pluripotent stem cells.

**STRENGTHS AND LIMITATIONS OF THIS STUDY:**

- We described a systematic, evidence-based approach towards drug repurposing in motor neuron disease (MND), specifically for Motor Neuron Disease – Systematic Multi-arm Adaptive Randomised Trial (MND-SMART), a phase III multi-arm multi-stage clinical trial in MND.
- Systematic reviews of clinical studies in neurodegenerative diseases and MND preclinical studies provided a robust evidence base to inform expert panel decisions on drug selection for clinical trials.
- Providing a contemporary evidence base using traditional systematic reviews is challenging given their time-consuming and labour-intensive nature.
- Incorporation of machine learning and automation tools for systematic reviews, and data from experimental drug screening can be helpful for future drug selection.

## INTRODUCTION

Motor neuron disease (MND), also known as amyotrophic lateral sclerosis (ALS), is a progressive neurodegenerative disease with a median survival of 2-3 years.[1] Despite many promising preclinical studies and 125 phase II and phase III trials reported between 2008 and 2019, riluzole remains the only globally approved disease-modifying treatment, prolonging survival by an average of two to three months.[2] Edavarone and masitinib have both emerged as potentially promising candidates in clinical trials, but treatment effects are modest and neither drug has received approval in Europe.[3, 4] In a long-term multi-centre prospective cohort study, edaravone showed no significant disease-modifying effect.[5] Previously, decisions to evaluate drugs in MND have been informed by preclinical studies, typically using mouse models, such as the SOD1^G93A^ mouse, despite known limitations in the extent to which such models recapitulate human pathology,[6] and concerns of the reproducibility of findings from such models.[7] Clinical trials in MND are further complicated by the challenges of designing and delivering trials in a rapidly progressive, heterogeneous, disabling and fatal disease with a lack of reliable and sensitive outcome measures or biomarkers.[2]

Over the same period there have, however, been rapid technical advances in MND genomics, human induced pluripotent stem cells and gene-editing, which have enabled better understanding of underlying pathophysiology (including potential shared pathways across neurodegenerative diseases), and the development of more sophisticated disease models. In parallel, drug repurposing (testing a drug already used or tested for other indications) has been successfully adopted in many diseases and can significantly reduce development time and cost, with the added benefit of the availability of prior safety data to guide selection.[8] In relapsing-remitting multiple sclerosis for instance, dimethyl fumarate, cladribine,[9], alemtuzumab,[10, 11] and rituximab[12] provide examples of successful repurposing as disease-modifying treatments.

Systematic review has been recommended to have a key role in planning new research studies.[13] We previously used a strategy based on systematic review to identify repurposed interventions for secondary progressive multiple sclerosis (MS). This involved a two-stage systematic review and meta-analysis assessing clinical and preclinical data to identify putative therapeutic interventions[14] and led to the Multiple Sclerosis – Secondary Progressive Multi-Arm Randomisation Trial (MS-SMART), a phase IIb multi-arm randomised controlled trial.[15, 16] The three drugs selected for MS-SMART were based in part on their availability for investigator-led clinical trials and did not show efficacy, but two of the top seven drugs thus identified, ibudilast (ranked first), and lipoic acid, have since shown promise in phase II studies in secondary progressive MS.[17, 18]

Noting similarities between MS and MND as neurodegenerative diseases with limited treatment options, in 2014 we embarked on a similar strategy to identify candidate oral neuroprotective agents in MND. In parallel, we developed the multi-arm multi-stage Motor Neuron Disease – Systematic Multi-Arm Adaptive Randomised Trial (MND-SMART, clinicaltrials.gov registration number: NCT04302870) to provide a more efficient pipeline to evaluate drugs in MND than conventional standalone two-arm trials.[19, 20] Here we describe the development and implementation of a systematic, structured, and unbiased evidence-based approach to inform the selection of potential oral neuroprotective agents for clinical evaluation in MND-SMART. Specifically, the purpose here is not to provide a contemporary summary of existing evidence, but to describe the process through which clinical trial drugs were selected.

## METHODS

The work was guided by a systematic review protocol. Over the duration of the project and given the novelty of this approach, this protocol was updated in the light of accumulating experience, and the complete record of the protocol, including the changes made, is available at Open Science Framework.[21]

### Overview

The overall drug selection strategy is characterised in Figure 1. We used systematic review to identify publications describing clinical trials or reports of the clinical use of drugs in MND and in four other neurodegenerative diseases which we considered might share pivotal pathways: Alzheimer’s disease (AD), Parkinson’s disease (PD), Huntington’s disease (HD) and MS. For MS, we excluded studies of relapsing-remitting disease since we were interested in drugs tested in the progressive phase where neurodegeneration is a major feature. We annotated publications for the drugs tested and diseases studied, taking forward drugs described in at least one MND publication or in publications in at least two other diseases. We scored each drug using a predefined framework evaluating efficacy, safety, study size and quality. In parallel, we performed a systematic review and meta-analysis of the preclinical MND and frontotemporal dementia (FTD; because of pathological overlap with MND) literature for these drugs. We summarised evidence from both reviews for each drug and presented these to an expert panel consisting of clinical and academic neurologists with expertise in MND, clinical trials, pharmacology/drug selection, and preclinical models of MND.

**Figure 1.**
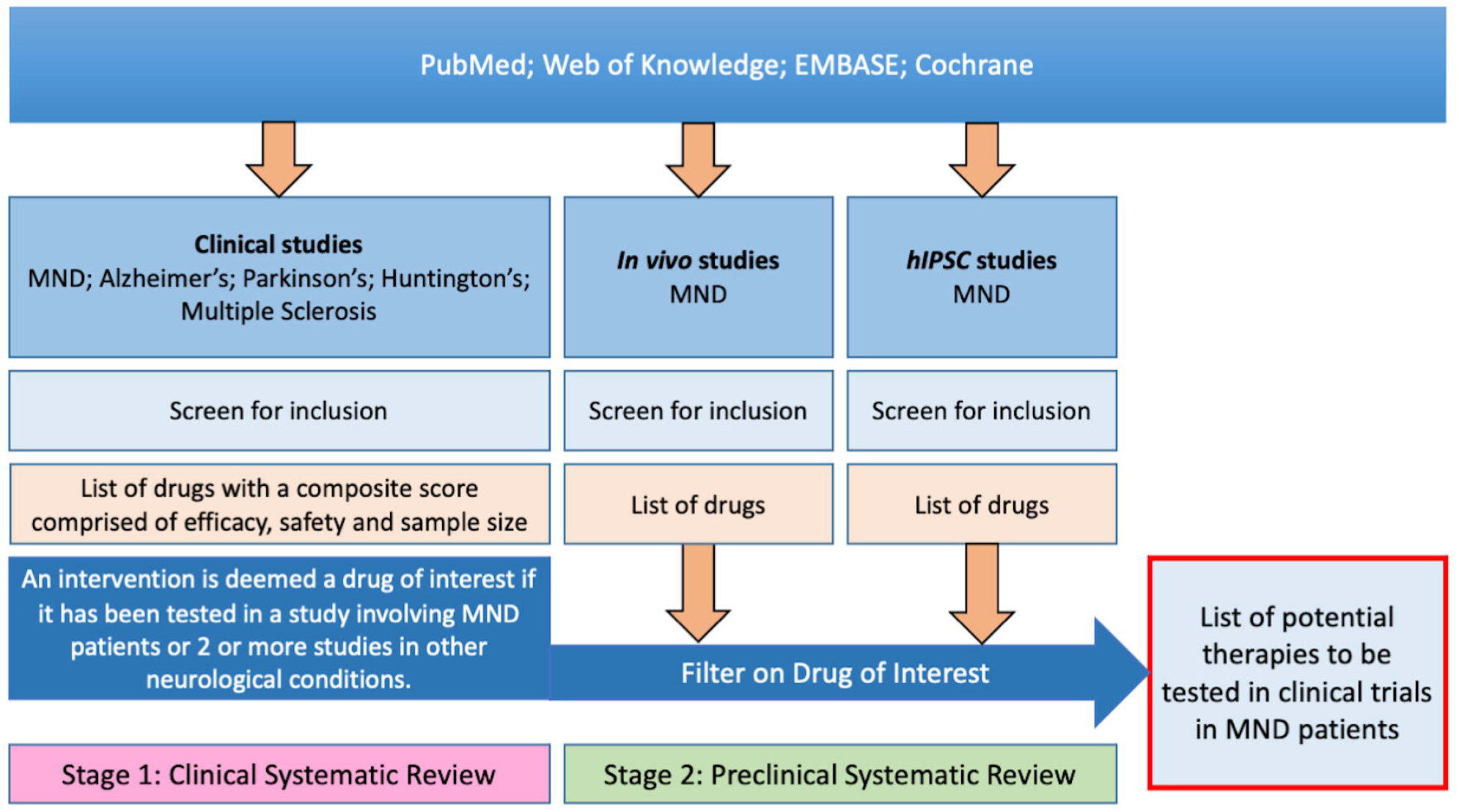
Diagram illustrating two-stage systematic review approach to inform identification and selection of putative treatments to take forward to clinical trial.

### Systematic review of clinical evidence

The MS-SMART drug selection process used the same strategy, except we selected drugs tested at least once in MS or in at least two other conditions. The protocol for and results from this search, conducted in September 2011, has been published.[14] That search involved three online databases (PubMed, ISI Web of Knowledge and EMBASE) using the terms “multiple sclerosis” OR “Alzheimer’s disease” OR “Huntington’s disease” OR “Parkinson’s disease” OR “motor neuron disease” OR “amyotrophic lateral sclerosis”. On 13 December 2013, we updated the search using the same terms but with limitations for PubMed to clinical trials, and date of record creation after 01/07/2011; for ISI Web of Knowledge to Document type “Clinical trial” and publication years: 2011, 2012 and 2013; for EMBASE: previous search string AND (“case series” or “case report” or “cohort study”), with limits: human studies, full text studies from 2011; and we also contacted the Cochrane Neuromuscular review group to obtain a list of interventions tested in MND/ALS. The protocol of this update was stored locally; in the light of increasing recognition of the importance of making systematic review protocols available, the protocol was published without amendment in September 2019.[22]

Two reviewers (MM and KE) independently screened title and abstracts of publications identified in the new search against the inclusion and exclusion criteria shown in Table 1**Error! Reference source not found**., with discrepancies resolved by discussion. We included case reports, uncontrolled case series, non-randomised parallel group studies, crossover studies, and randomised controlled trials with any report of safety or efficacy. We extracted basic information from each publication including author, year of publication, intervention tested and disease.

**Table 1:** Eligibility criteria for clinical systematic review.

|  |
| --- |
| **Inclusion criteria** |
| - Publications reporting qualitative or quantitative data provided on either safety or efficacy of an orally delivered intervention in people with motor neuron disease / amyotrophic lateral sclerosis, Alzheimer's disease, Parkinson's disease, Huntington's disease or multiple sclerosis<br>- Studies reporting change in clinical status (including death, tracheostomy free survival, relapse frequency, disability progression, behavioural symptoms), or changes in biomarkers (including magnetic resonance imaging, blood, cerebrospinal fluid and muscle strength) |
| **Exclusion criteria** |
| - Isolated reporting of non-pharmacological interventions such as acupuncture, aromatherapy, physiotherapy or exercise<br>- Articles reporting the use of interventions already licensed for clinical use in MND such as riluzole<br>- Articles on levodopa treatment for Parkinson's disease<br>- Studies reporting different modes of intervention delivery other than oral administration<br>- Publications reporting secondary analysis of previously published clinical trial data<br>- Protocols for clinical trials<br>- Preventative studies<br>- Reviews<br>- Studies on healthy volunteers<br>- Studies in patients with relapsing-remitting MS<br>- Studies reporting combination treatments including where an oral and a non-oral intervention are administered<br>- Publications where disease type is not specified to be in keeping with the included diseases (studies of vascular dementia, mild cognitive impairment and dementias other than Alzheimer's disease are excluded)<br>- Studies on patients with parkinsonism are excluded as this do not imply Parkinson's disease exclusively)<br>- Publications describing studies where multiple drugs were tested in a cohort without any data on individual drugs |

For all candidate interventions which had not been excluded based on feasibility or plausibility, we extracted further information on safety, efficacy, quality of study and study size from publications to a Microsoft Access database and scored these against a predefined metric (Tables 2 and 3). For each drug, we calculated an overall drug score by taking the product of the mean score in each domain for safety, efficacy, quality, study size and multiplying this by log_10_(1+number of publications). We then ranked drugs according to these scores.

**Table 2:** Scoring metric for clinical review

|  |
| --- |
| **Safety Score [S]**<br>“not described”: 1 point<br>“SUSARs (Suspected Unexpected Serious Adverse Reactions) or mortality observed”: 1 point<br>“SAEs (Serious Adverse Events) only”: 2 points<br>“AEs (Adverse events) only”: 3 points<br>“No adverse effects reported”: 4 points |
| **Efficacy Score [E]**<br>***Efficacy score is assigned based on primary outcome measure, and where this is not identified, on the mean efficacy score for all outcomes reported in each publication.***<br>“not presented”: 1 point<br>“definite (i.e. statistically significant) worsening”: 1 point<br>“neutral”: 2 points<br>“non-significant improvement”: 3 points<br>“significant improvement”: 4 points |
| **Quality Score [Q]**<br>Study quality was assessed using a combination of criteria taken from a risk of bias tool developed through a Delphi process, GRADE and CAMARADES methods as shown in Table 3. Once each publication has been scored they are sorted in quartiles of study quality based on the total number of checklist items scored, with the lowest quartile scoring 1 point and the highest quartile scoring 4 points. |
| **Study size score [SS]**<br>“1-10 patients”: 1 point<br>“11-100 patients”: 2 points<br>“101-1000 patients”: 3 points<br>“>1000 patients”: 4 points |

**Table 3:** Scoring method for evaluation of study quality in clinical systematic review.

|  | CAMARADES | Delphi | GRADE |
| --- | --- | --- | --- |
| **Binary response items :**<br>*Yes (1 point); No (0 points)* |  |  |  |
| Peer reviewed publication | X |  |  |
| Statement of potential conflicts of interest | X |  |  |
| Sample size calculation | X | X |  |
| Random allocation to group | X | X | X |
| Allocation concealment | X |  | X |
| Blinded assessment of outcome |  | X |  |
| **Tertiary response items:**<br>*Yes (1 point); No (0 points); Not clear (0.5 points)* |  |  |  |
| Were the groups similar at baseline regarding the most important prognostic indicators? |  | X |  |
| Were the eligibility criteria specified? |  | X |  |
| Were point estimates and measures of variability presented for the primary outcome measures? |  | X |  |
| Was there intention to treat analysis? |  | X |  |
| Complete accounting of patient and outcome events |  |  | X |
| Non-selective outcome reporting |  | X |  |
| No other limitations |  |  | X |
| Can we be confident in the assessment of outcome? |  |  | X |
| **Quinary response items:**<br>*N/A; Definitely yes (1 point); Probably yes (0.75 points); Probably no (0.25 points); Definitely No (0 points)* |  |  |  |
| Was selection of treatment and control groups drawn from the same population? |  |  | X |
| Can we be confident that patients received the allocated treatment? |  |  | X |
| Can we be confident that the outcome of interest was not present at start of the study? |  |  | X |
| Did the study stratify on variables associated with the outcome of interest or did the analysis take this into account? |  |  | X |
| Can we be confident in the assessment of the presence or absence of prognostic factors? |  |  | X |
| Was the follow up of cohorts adequate? |  |  | X |
| Were co-interventions similar between groups? |  |  | X |

### Systematic review of preclinical evidence

In parallel, we performed a systematic review of the preclinical literature (date of search 6^th^ April 2016), focussing on publications describing the candidate interventions which had not been excluded on the basis of feasibility or plausibility, and using our previously published systematic review protocol.[23] We evaluated data from all *in vivo* models of MND and FTD including (i) mammalian models (mouse and rat), (ii) organisms with a central nervous system (*Drosophila, Caenorhabditis elegans* and Zebrafish) and (iii) multicellular eukaryotic models such as yeast. We also include data from studies using human induced pluripotent stem cells (iPSCs) derived from people with MND.

### Patient and public involvement

The MND-SMART group has consulted people with MND, their families and carers via a patient and public involvement advisory group throughout the development of the trial. The group recommended using liquid investigational medicinal product formulations to enable patients with swallowing difficulties including those using gastrostomy to participate and remain in the trial. This was taken into consideration by the expert panel during the drug selection process.

### Expert panel review

A panel of six independent experts consisting of neurologists with expertise in translational neurology, clinical MND research, meta-analysis and clinical trials reviewed all data. The panel rated drugs as “green” (most favourable), “amber” (less favourable) and “red” (least favourable) based on biological plausibility; safety profile; and ranking by drug score from clinical literature. Drugs rated “red” for any criteria were excluded, along with drugs which had been tested in more than three previous trials in MND. Next, the panel ranked shortlisted drugs considering also preclinical evidence and practical issues such as availability in oral liquid formulation, and restricted availability without prescription (to discourage off-protocol self-medication from study participants). As this approach might not cover novel drugs or pathways that have yet to be tested clinically in neurodegenerative diseases, the panel were given flexibility to consider emerging evidence for hitherto unconsidered drugs.

## RESULTS

### Clinical systematic review and initial screening of candidate interventions

The PRISMA diagram for the clinical review is shown in Figure 2. Literature search in August 2011 of PubMed, ISI Web of knowledge and EMBASE, and Cochrane list of clinical trials in MS for MS-SMART identified 29500 publications. 12893 duplicates were removed, and 15232 publications did not meet the inclusion criteria. 1375 publications were included from this initial search.

**Figure 2.**
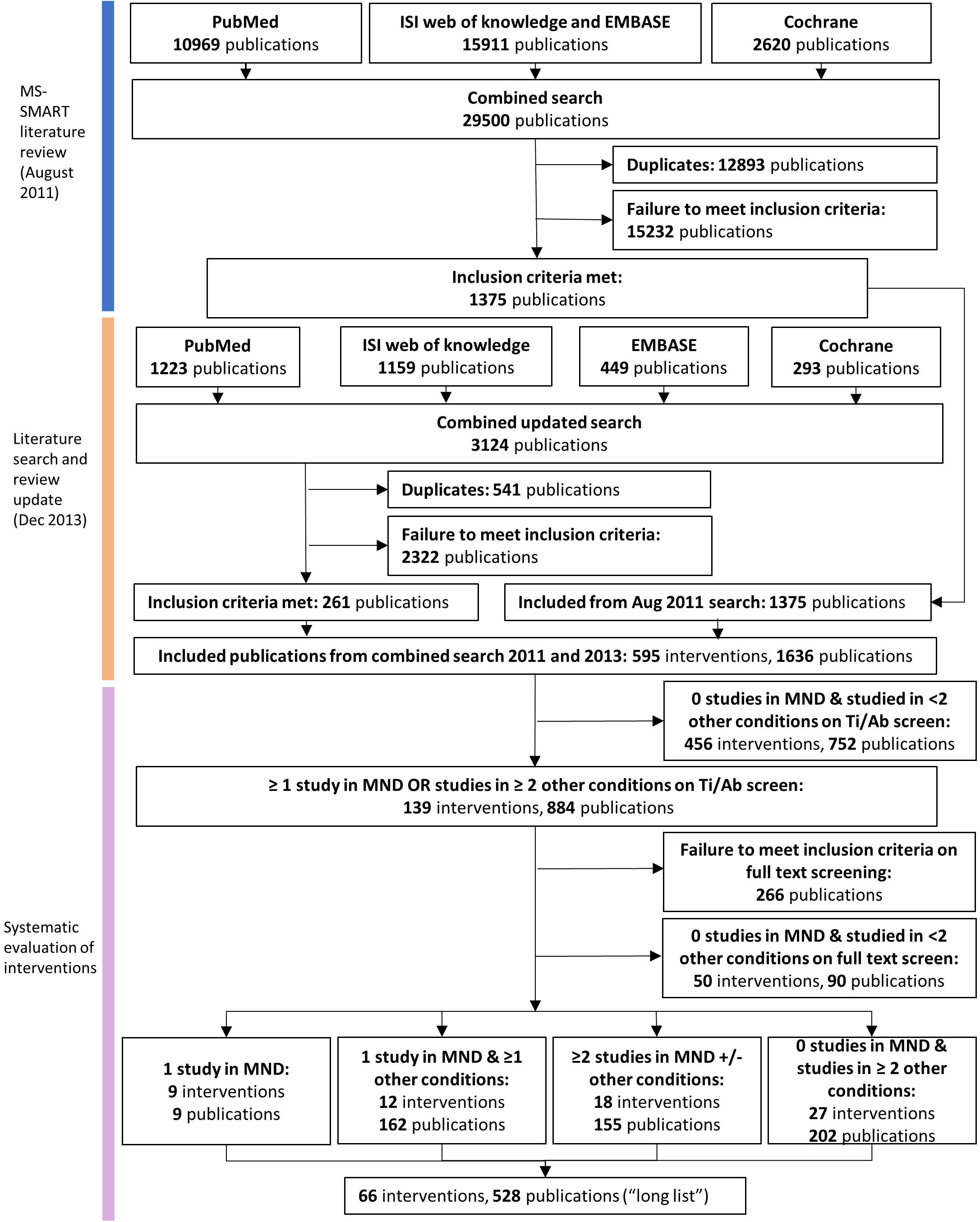
PRISMA diagram for clinical systematic review. Ti/Ab: Title/Abstract

In the updated search in December 2013 a further 3124 publications were identified from PubMed, ISI Web of Knowledge, EMBASE and Cochrane databases. 541 duplicates were removed, and 2322 publications did not meet the inclusion criteria. 261 publications were included.

Based on information contained in the Title and Abstract of these 1636 included publications we identified 595 interventions, of which 139 met our criteria of being described in at least one MND publication or in publications in two other diseases, in a total of 884 publications. On full text screening, 266 of these 884 publications did not meet our inclusion criteria.A further 50 interventions described in 90 publications were were excluded because it became apparent at full text screening that the intervention has not been tested either in MND or in at least two of the other diseases.. The remaining 66 interventions (528 publications) were scored against our predefined criteria and ranked (Table 4). During preparation of this manuscript, we discovered that a publication describing the effect of N-acetyl cysteine in MND had been included in error, as no data were available for N-acetyl cysteine monotherapy.[24]

**Table 4:** Longlisted interventions ranked by drug score from clinical review

| Intervention | Number of Publications | Quality Score | Efficacy Score | Safety Score | Study Size Score | Drug score |
| --- | --- | --- | --- | --- | --- | --- |
| Rivastigmine | 29 | 3.34 | 3.33 | 2.1 | 2.62 | 90.62 |
| Memantine | 51 | 3.02 | 2.87 | 2.2 | 2.47 | 80.7 |
| Vitamin D3 | 11 | 3.27 | 3.01 | 3.36 | 2.18 | 78.08 |
| Donepezil | 41 | 3.1 | 2.72 | 2.51 | 2.24 | 76.99 |
| Pramipexole | 14 | 3.07 | 3.04 | 2.43 | 2.64 | 70.41 |
| Galantamine | 12 | 2.75 | 2.85 | 2.67 | 2.5 | 58.29 |
| Amantadine | 59 | 2.37 | 3 | 2.36 | 1.9 | 56.53 |
| Dextromethorphan/<br>quinidine | 3 | 4 | 3.33 | 2.33 | 3 | 56.19 |
| Selegiline | 21 | 3 | 2.59 | 2.1 | 2.38 | 51.98 |
| 4-Aminopyridine | 10 | 3.2 | 2.76 | 2.5 | 2.1 | 48.28 |
| Acetyl-L-Carnitine | 10 | 2.9 | 2.64 | 2.5 | 2.3 | 45.83 |
| Simvastatin | 5 | 3.6 | 2.27 | 2.8 | 2.4 | 42.73 |
| Lamotrigine | 6 | 4 | 2.25 | 2.33 | 2.33 | 41.41 |
| Bromocriptine | 13 | 2.92 | 2.4 | 2.54 | 1.92 | 39.21 |
| Clozapine | 6 | 3 | 2.9 | 2.5 | 2 | 36.8 |
| Gabapentin | 9 | 2.67 | 2.4 | 2.44 | 2.33 | 36.46 |
| Creatine | 12 | 2.67 | 2.14 | 2.33 | 2.42 | 35.87 |
| Ginkgo biloba | 3 | 3.67 | 2.61 | 3.67 | 1.67 | 35.21 |
| Minocycline | 11 | 2.45 | 2.27 | 2.64 | 2.18 | 34.54 |
| Vitamin E | 9 | 3.33 | 2.1 | 2 | 2.44 | 34.25 |
| Levetiracetam | 6 | 3 | 3.33 | 2.17 | 1.83 | 33.57 |
| Atomoxetine | 4 | 3.25 | 2.5 | 3.25 | 1.75 | 32.3 |
| Coenzyme Q10 | 9 | 3.33 | 2.16 | 1.89 | 2.33 | 31.67 |
| Tacrine | 10 | 3.3 | 2.81 | 1.8 | 1.8 | 31.26 |
| Olanzapine | 5 | 2.4 | 3.47 | 2.6 | 1.6 | 26.93 |
| Estrogen | 9 | 3 | 2.66 | 1.67 | 2 | 26.57 |
| Nimodipine | 5 | 3 | 2.35 | 2.2 | 2.2 | 26.55 |
| Riluzole | 17 | 2.41 | 2.35 | 1.76 | 2.06 | 25.88 |
| Ciclosporin | 9 | 2.78 | 1.93 | 2.11 | 2.22 | 25.15 |
| Dextromethorphan | 7 | 2.29 | 2.4 | 2.71 | 1.86 | 24.97 |
| Naltrexone | 8 | 2.5 | 2.22 | 2.62 | 1.75 | 24.29 |
| Theophylline | 2 | 4 | 3.25 | 2.5 | 1.5 | 23.26 |
| Valproate | 9 | 2.56 | 2.33 | 2 | 1.89 | 22.53 |
| Fluoxetine | 6 | 2.67 | 2.42 | 2.33 | 1.67 | 21.18 |
| Levamisole | 3 | 2.67 | 2.78 | 2.33 | 2 | 20.81 |
| Melatonin | 8 | 2 | 2.11 | 2.75 | 1.88 | 20.81 |
| Celecoxib | 2 | 4 | 2 | 1.5 | 3 | 17.18 |
| 3,4-Diaminopyridine | 6 | 2.83 | 2.4 | 2.17 | 1.33 | 16.62 |
| Milacemide | 4 | 3 | 1.75 | 2.25 | 2 | 16.51 |
| N-acetyl cystine | 1 | 3 | 3 | 3 | 2 | 16.26 |
| Tranylcypromine | 2 | 2.5 | 2.5 | 3.5 | 1.5 | 15.66 |
| Aspirin | 4 | 2.75 | 2 | 1.75 | 2.25 | 15.14 |
| Ursodeoxycholic acid | 1 | 4 | 2 | 3 | 2 | 14.45 |
| Tolbutamide | 2 | 3 | 2.5 | 2 | 2 | 14.31 |
| Imipramine | 2 | 3.5 | 2 | 2 | 2 | 13.36 |
| Lithium | 12 | 2.42 | 2.19 | 1.5 | 1.5 | 13.29 |
| Modafinil | 2 | 4 | 3.33 | 1 | 2 | 12.72 |
| Omega 3 Fatty Acid | 2 | 2.5 | 1.75 | 3 | 2 | 12.52 |
| Octacosanol | 2 | 2.5 | 2 | 3.5 | 1.5 | 12.52 |
| Indinavir | 2 | 3.5 | 1.8 | 1.5 | 2.5 | 11.27 |
| Sodium phenylbutyrate | 1 | 4 | 2 | 2 | 2 | 9.63 |
| Tilorone | 1 | 4 | 2 | 2 | 2 | 9.63 |
| lipoic acid | 2 | 2.5 | 2 | 2 | 2 | 9.54 |
| Isoprinosine | 4 | 3 | 1.75 | 1.25 | 2 | 9.17 |
| Tetrahydrocannabinol | 2 | 3.5 | 1.5 | 1.5 | 2 | 7.51 |
| Topiramate | 1 | 4 | 1 | 2 | 3 | 7.22 |
| Haloperidol | 2 | 3 | 2.33 | 1 | 1.5 | 5.01 |
| Amino acid mixture | 5 | 1.6 | 2 | 1 | 2 | 4.98 |
| Rolipram | 2 | 1.5 | 2 | 3 | 1 | 4.29 |
| Alsamin | 1 | 2 | 3.5 | 1 | 2 | 4.21 |
| Pentoxifylline | 3 | 2 | 1.61 | 1 | 2 | 3.88 |
| Verapamil | 1 | 2 | 2 | 1 | 2 | 2.41 |
| IGF-1 | 1 | 2 | 1 | 1 | 3 | 1.81 |
| Propranolol | 1 | 1 | 3 | 1 | 2 | 1.81 |
| Fluvoxamine | 2 | 1 | 3 | 1 | 1 | 1.43 |
| Amitriptyline | 1 | 1 | 1 | 3 | 1 | 0.9 |

### Preclinical systematic review

We identified 14195 publications. After removing duplicates, two independent researchers screened Title and Abstract of 7586 unique publications, with differences reconciled by a third reviewer. 396 studies were included. 330 studies reported effects on survival of the model organism and were included in the meta-analysis. Of the 66 longlisted interventions from the clinical review, 20 had been tested in in vivo models reporting an endpoint of survival (Table 5), and of these 17 had at least one report of improved survival. Because survival was reported in a single paper for 11 of these 20 interventions, we did not conduct a meta-analysis.

**Table 5:** Summary of preclinical studies evaluating the effect of interventions longlisted from the clinical review on survival outcomes. All listed studies used mouse models. LogMSR = log(median survival in treatment group/median survival in control group)

| Publication | Drug | Total number of animals | Median survival in treatment group | Median survival in control group | logMSR |
| --- | --- | --- | --- | --- | --- |
| Kira 2006 | Acetyl-L-Carnitine | 20 | 270 | 240 | 0.1178 |
| Barneoud 1999 | Aspirin | 38 | 150 | 155 | -0.0328 |
| Tanaka 2011 | Bromocriptine | 69 | 40 | 35 | 0.1335 |
| Drachman 2002 | Celecoxib | 55 | 139 | 119 | 0.1554 |
| Karlsson 2004 | Ciclosporin | 13 | 144 | 130 | 0.1023 |
| Keep 2001 | Ciclosporin | 11 | 24 | 12 | 0.6931 |
| Turner 2003 | Clozapine | 16 | 140 | 132 | 0.0588 |
| Andreassen 2001 | Creatine | 24 | 155 | 135 | 0.1382 |
| Kaddurah-Daouk 2000 | Creatine | 13 | 169 | 144 | 0.1601 |
| Klivenyi 2004 | Creatine | 22 | 150 | 125 | 0.1823 |
| Choi 2008 | Estrogen | 70 | 135 | 127 | 0.0611 |
| Koschnitzky 2014 | Fluoxetine | 34 | 139 | 132 | 0.0517 |
| Gurney 1996 | Gabapentin | 17 | 140 | 139 | 0.0072 |
| Gurney 1996 | Gabapentin | 38 | 175 | 165 | 0.0588 |
| Ferrante 2001 | Ginkgo Biloba | 20 | 136 | 125 | 0.0843 |
| Fornai 2008 | Lithium | 20 | 146 | 117 | 0.2214 |
| Gill 2009 | Lithium | 55 | 124 | 127 | -0.0239 |
| Pizzasegola 2009 | Lithium | 20 | 119 | 129 | -0.0807 |
| Dardiotis 2013 | Melatonin | 28 | 143 | 143 | 0.0000 |
| Weishaupt 2006 | Melatonin | 50 | 137 | 131 | 0.0448 |
| Zhang 2013 | Melatonin | 30 | 145 | 137 | 0.0568 |
| Wang 2005 | Memantine | 21 | 130 | 122 | 0.0635 |
| Keller 2011 | Minocycline | 32 | 147 | 138 | 0.0632 |
| Kriz 2002 | Minocycline | 29 | 364 | 336 | 0.0800 |
| Van Den Bosch 2002 | Minocycline | 14 | 155 | 130 | 0.1759 |
| Zhang 2003 | Minocycline | 20 | 140 | 130 | 0.0741 |
| Zhu 2002 | Minocycline | 20 | 135 | 127 | 0.0611 |
| Andreassen 2000 | N-acetyl cysteine | 30 | 134 | 129 | 0.0380 |
| Jaarsma 1998 | N-acetyl cysteine | 28 | 251 | 239 | 0.0490 |
| Yip 2013 | Omega 3 | 32 | 182 | 182 | 0.0000 |
| Petri 2006 | Sodium Phenylbutyrate | 26 | 139 | 127 | 0.0903 |
| Ryu 2005 | Sodium Phenylbutyrate | 40 | 145 | 127 | 0.1325 |
| Crochemore 2009 | Valproate | 11 | 140 | 140 | 0.0000 |
| Rouaux 2007 | Valproate | 36 | 115 | 110 | 0.0445 |
| Sugai 2004 | Valproate | 17 | 295 | 265 | 0.1072 |
| Gianfocarò 2013 | Vitamin D | 100 | 126 | 124 | 0.0160 |

### Shortlisted candidate drugs for clinical trial

The expert panel met in 2017 and considered the clinical and preclinical research summaries for each of these 66 drugs, shortlisting 22 for further consideration. An evidence summary was compiled for each of these drugs including the following information: (1) If they had been tested in 3 or more *in vivo* MND studies, (2) the number of clinical trials in people with MND, (3) the putative target pathway, (4) feasibility for delivery via enteral tube (noting that swallowing is commonly affected in MND), (5) detailed safety information including common side effects, rare but serious side effects and requirements for monitoring, (6) published clinical studies in MND, and (7) clinical trials registered on clinicaltrials.gov. The expert panel met on 9^th^ January 2017 and discussed the evidence for each drug. 11 drugs were excluded in the first round. Following a second round of discussions, four other drugs were excluded. Reasons for exclusion are detailed in Table 6.

**Table 6:**
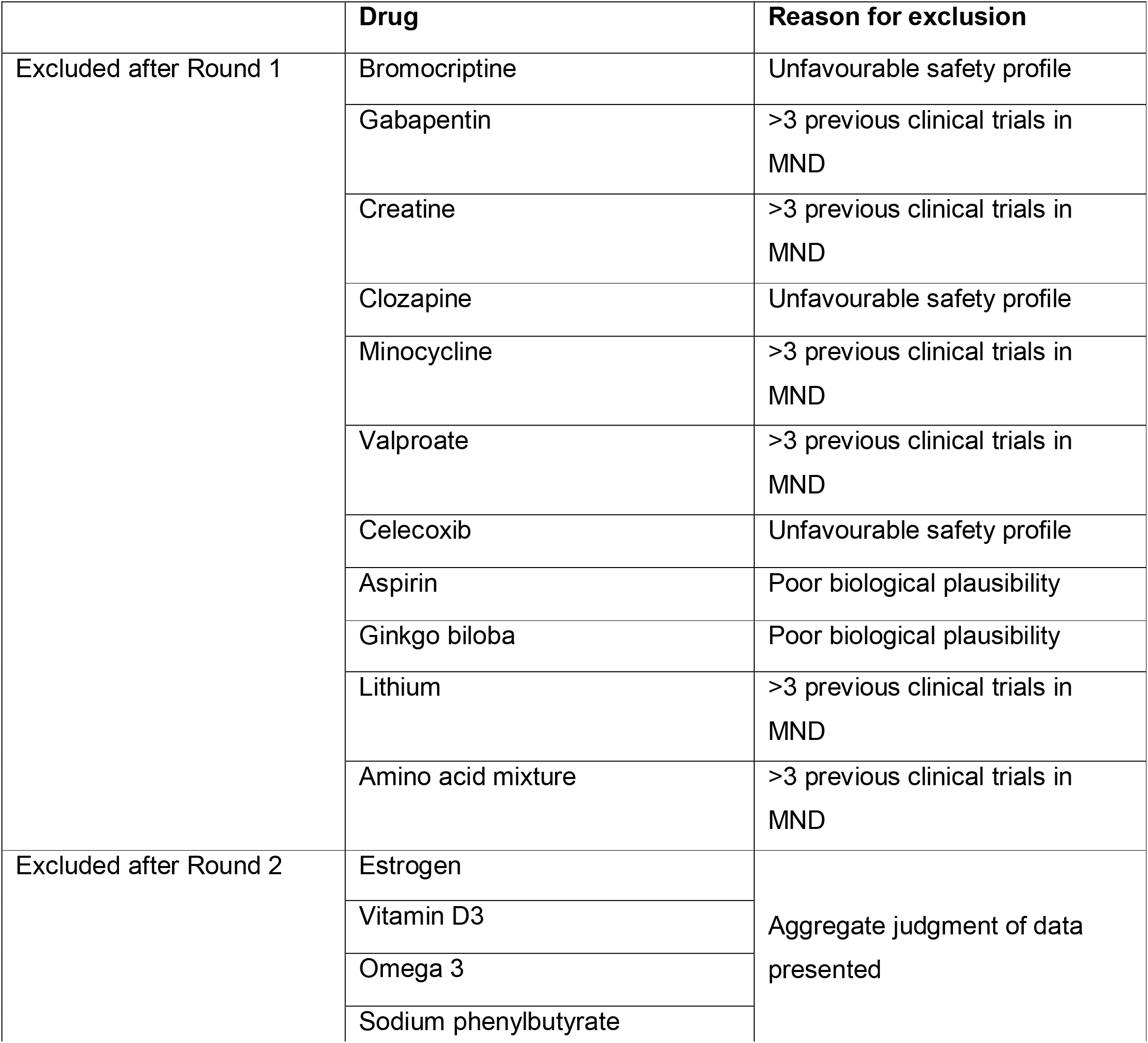
Drugs excluded following expert panel review and reasons for exclusion.

The seven candidate drugs remaining were memantine, acetyl-l-carnitine, simvastatin, ciclosporin, melatonin, fluoxetine and N-acetyl cysteine. The evidence for each final shortlisted drug is summarised in Tables 7 and 8. The panel also considered emerging evidence relating to trazodone, and following due consideration recommended memantine and trazodone as the first two investigational medicinal products for MND-SMART.

**Table 7:** Number of publications of clinical studies and participants according to type of disease for the final seven shortlisted drugs. AD: Alzheimer’s disease; HD: Huntington’s disease; MND: Motor neuron disease; MS: Multiple sclerosis; PD: Parkinson’s disease.

| Drug | Number of publications |  |  |  |  |  | Number of participants |  |  |  |  |  |
| --- | --- | --- | --- | --- | --- | --- | --- | --- | --- | --- | --- | --- |
|  | MND | AD | HD | MS | PD | Total | MND | AD | HD | MS | PD | Total |
| Acetyl-L-carnitine | 0 | 9 | 1 | 0 | 0 | 10 | 0 | 1224 | 10 | 0 | 0 | 1234 |
| Ciclosporin | 2 | 0 | 0 | 7 | 0 | 9 | 110 | 0 | 0 | 1092 | 0 | 1202 |
| Fluoxetine | 0 | 0 | 1 | 2 | 3 | 6 | 0 | 0 | 30 | 51 | 32 | 113 |
| Melatonin | 1 | 4 | 0 | 0 | 3 | 8 | 3 | 273 | 0 | 0 | 64 | 340 |
| Memantine | 1 | 32 | 2 | 1 | 15 | 51 | 63 | 11912 | 39 | 116 | 809 | 12939 |
| N-acetyl cysteine | 0 | 1 | 0 | 0 | 0 | 1 | 0 | 47 | 0 | 0 | 0 | 47 |
| Simvastatin | 0 | 3 | 0 | 1 | 1 | 5 | 0 | 469 | 0 | 307 | 12 | 788 |

**Table 8:**
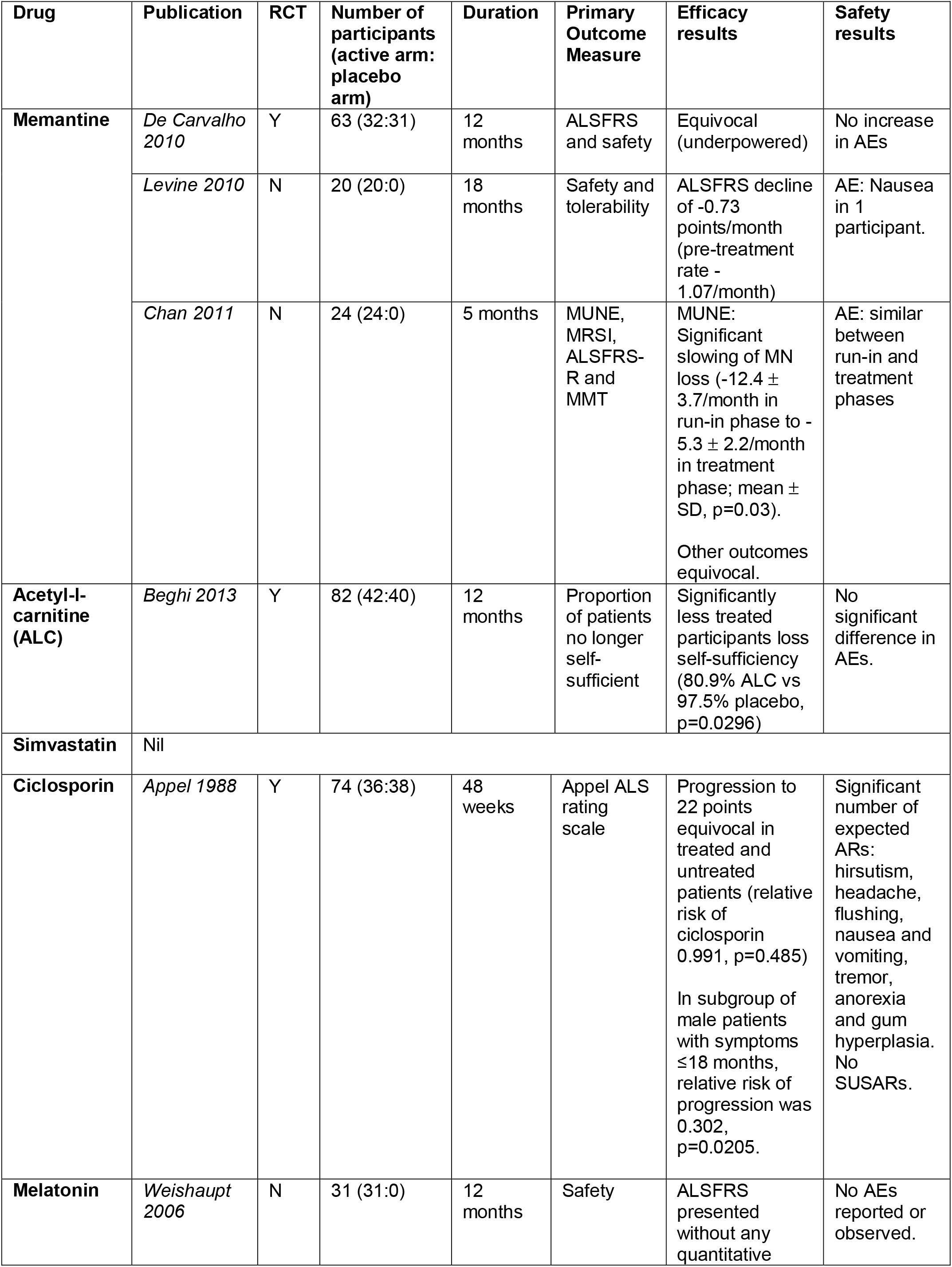
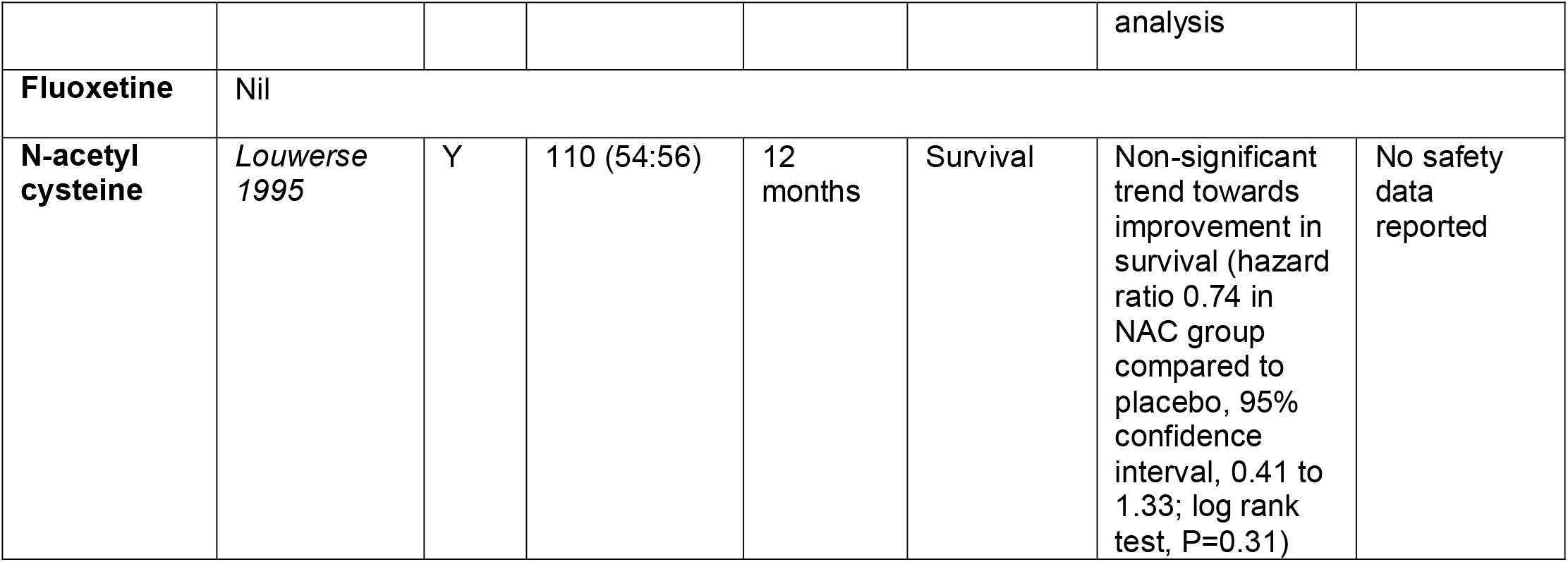
Summary of previous MND clinical trials for final shortlisted drugs. RCT: Randomised controlled trial; N: number of participants. AE: Adverse events; AR: Adverse reactions; ALC: Acetyl-l-carnitine; ALSFRS: Amyotrophic Lateral Sclerosis Functional Rating Scale; MMT: Manual Muscle Testing; MN: Motor neuron; MRSI: Magnetic Resonance Spectroscopy Imaging; MUNE: Motor unit number estimate; NAC: N-acetyl cysteine; SD: Standard deviation; SUSAR: Suspected Unexpected Severe Adverse Reaction.

## DISCUSSION

Since drugs have undergone rigorous safety and pharmacokinetic testing, drug repurposing –– the use of an established drug in a novel therapeutic indication –– reduces costs and barriers to clinical development. Our experience of the successful application of a systematic approach to selecting neuroprotective drugs for repurposing in MS clinical trials[14] encouraged us to use a similar approach in MND. The first part of the review assessed clinical data in MND and in other neurodegenerative diseases with potential shared pathophysiological pathways. This allowed for the identification of drugs with good central nervous system penetrance and the potential for efficacy and safety in patients with neurodegenerative diseases. However, drug selection based on clinical data alone is biased towards those tested in conditions where large well-designed randomised controlled trials have been performed and where the mechanism of action may be particular to that condition. Notably, two of our top five ranked drugs were cholinesterase inhibitors licensed for Alzheimer’s disease, a mechanism less relevant to MND. It was therefore important that we augment this approach with expert opinion and with preclinical data in MND and frontotemporal dementia models to provide mechanistic relevance. Taken together we have compiled evidence from clinical and preclinical data and used this to inform the selection of potential oral neuroprotective agents for clinical evaluation in people with MND. Through sequential systematic review and meta-analysis, we identified a short list of 22 candidate interventions selected from an initial set of 595 drugs.

While some identified drugs demonstrate a good safety profile and have a relevant putative target pathway in MND, others have less favourable side effects profiles or a requirement for close therapeutic monitoring (e.g. clozapine) which necessitates a higher threshold of evidence before testing in clinical trial. This highlights another advantage of our approach, in that it allows the identification of interventions that warrant further rigorous preclinical testing (“cislation”[25]) in *in vivo* or *in vitro* models of ALS, with a view to providing more robust information for efficacy to support their inclusion in future clinical trials.

Following rounds of discussion, the expert panel identified memantine as a drug to be tested in MND-SMART. Memantine is a non-competitive N-methyl-D-aspartate (NMDA) receptor antagonist used in the treatment of moderate to severe Alzheimer’s disease. It was shown to significantly delay disease progression and improve survival in mouse models carrying a high copy number of SOD1^G93A^. [26] Memantine has been previously tested in three MND clinical trials. A phase II double-blind placebo-controlled study of 63 patients powered to evaluate safety and tolerability did not identify any increase in adverse events.[27] There was a trend towards improvement in patients treated with memantine 20 mg/day, but no significant difference in ALSFRS-R. In a 5-month randomised double-blind study of 24 participants with ALS, there was significant slowing of spinal motor neuron loss as demonstrated on motor unit estimation testing in the high dose group (10 mg twice daily) compared to low dose (5 mg twice daily).[28] Adverse events were not reported. In a single-arm pilot study of 19 ALS patients, patients treated with riluzole and memantine had reduction in rate of ALSFRS decline and reduced CSF tau levels without any increase in adverse events.[29]

We also asked the expert panel to consider other drugs for which relevant data had only become available after the searches described here had been performed. Trazodone was nominated for consideration through this route. Trazodone is an atypical serotonin antagonist and reuptake inhibitor antidepressant. An unbiased drug screen found that trazodone inhibited Protein Kinase RNA-like endoplasmic reticulum kinase (PERK), which is pivotal to stress granule formation, a common feature of neurodegenerative diseases.[30] Inhibition of PERK was found to be beneficial in a fly model of ALS as well as in an *in vitro* neuronal assay of TDP-43 injury.[31] Furthermore, trazodone has been shown to modulate the ER-stress response resulting in an improvement in survival in animal models of prion disease and FTD.[30] and to be involved in mitochondrial energy metabolism and fatty acid synthesis in animal models of Huntington’s disease, and may prevent mitochondrial dysfunction in MND.[32] In a randomized double-blind placebo-controlled crossover phase II trial in 31 patients with FTD, trazodone was found to improve cognition as assessed by the neuropsychiatric inventory.[33] In trials of trazodone in Parkinson’s and Alzheimer’s disease, although there was no improvement in cognition, symptoms of sleep disturbance and depression were alleviated and adverse events were not increased.[34, 35]

### Limitations of this approach

The main challenge in this approach to drug selection is the ambition to base drug choice on the most contemporary evidence. Systematic reviews are time consuming, as evidenced by the interval between our updated search (2013) and expert committee consideration (2017). Furthermore, drugs with promising data in some domains would be excluded if they have been tested in only one disease other than MND; or if they have not been tested clinically despite overwhelming preclinical evidence. We excluded combination therapies, but it may be – as in the treatment of various cancers[36] and infections[37, 38] – that engagement with multiple targets is required to achieve a substantial disease-modifying effect.

Finally, some have claimed that the literature-based systematic review approach to drug selection is intrinsically flawed because it does not take into account disease specific pathophysiology (which may be largely unknown).[39] While the three drugs tested in MS-SMART were not effective,[15] we note that two other drugs on the final MS-SMART shortlist - ibudilast[17] and lipoic acid[18] -have since shown promise in independent phase II trials. Lipoic acid has been identified again as a favourable candidate drug in a further, independent review in 2020.[40] We sought to address this issue here by considering, in addition to clinical information, data from in vivo and in vitro research. Although much successful drug repurposing has been opportunistic and serendipitous, we recognise that future efforts should include consideration of our mechanistic understanding of neurodegenerative diseases and should systemically incorporate additional target and pathway-based information.[8]

### Future approaches to drug selection in MND-SMART

Ongoing rounds of drug selection for MND-SMART exploit innovations in automating literature searches, screening, and annotation, with these algorithms trained using the human efforts in the work reported here. These techniques show substantial improvements in efficiency in other fields.[41] Using the Systematic Review Facility (SyRF) (https://syrf.org.uk)[42] we have enabled a ‘living’ systematic review with automatic search, citation screening, identification of disease and drug, and selection of drugs meeting our criteria for the range of diseases in which studies have been performed. Because of similarities between MND and frontotemporal dementia we have included this as an additional disease of interest. Further details are extracted from full text publications of shortlisted drugs by a combination of machine and human work enabled through the SyRF platform, with human monitoring of machine decisions. The incorporation of machine learning and text mining techniques substantially reduces the human effort required and makes this approach feasible in the context of timely drug selection for adaptive clinical trials.

Complementing our literature-based approach, our current platform incorporates data from additional domains, including in house *in vitro* high throughput screening using human induced pluripotent stem cell culture; pathway and network analysis; and mining of drug and trial databases. We have also sought a broader range of inputs to our expert committee such that it now includes those with experience and expertise in managing people with MND and their symptoms, and of clinical trials, translational and clinical neurology, systematic reviews, experimental drug screening, pharmacology, chemistry, and drug discovery.

## CONCLUSIONS

We describe our experience in conducting a systematic, structured, unbiased and evidence-based approach to the selection of candidate drugs for evaluation in a clinical trial in MND by combining review of clinical and preclinical literature, and expert panel input. The first two drugs selected are memantine and trazodone. For future selection, we will incorporate machine learning and text mining to our systematic reviews and data from our drug discovery platform.

## Data Availability

Data are available in the Supplementary material.

## CONTRIBUTIONS

### Project management

Suvankar Pal, Siddharthan Chandran, and Malcolm R. Macleod. **Project administration:** Charis Wong, Jenna M. Gregory, Jing Liao, Kieren Egan, Hanna M. Vesterinen, and Malcolm R. Macleod.

### Conceptualisation

Jenna M. Gregory, Kieren Egan, Hanna M. Vesterinen, Siddharthan Chandran, and Malcolm R. Macleod.

### Systematic searching, screening, annotation and data extraction

Charis Wong, Jenna M. Gregory, Jing Liao, Kieren Egan, Hanna M. Vesterinen, Aimal Ahmad Khan, Maarij Anwar, Caitlin Beagan, Fraser Brown, John Cafferkey, Alessandra Cardinali, Jane Yi Chiam, Claire Chiang, Victoria Collins, Joyce Dormido, Elizabeth Elliott, Peter Foley, Yu Cheng Foo, Lily Fulton-Humble, Angus B. Gane, Stella A. Glasmacher, Áine Heffernan, Kiran Jayaprakash, Nimesh Jayasuriya, Amina Kaddouri, Jamie Kiernan, Gavin Langlands, Danielle Leighton, Jiaming Liu, James Lyon, Arpan R. Mehta, Alyssa Meng, Vivienne Nguyen, Na Hyun Park, Suzanne Quigley, Yousuf Rashid, Andrea Salzinger, Bethany Shiell, Ankur Singh, Tim Soane, Alexandra Thompson, Olaf Tomala, Fergal M. Waldron, Suvankar Pal, and Malcolm R. Macleod.

### Data curation

Charis Wong, Jenna M. Gregory, Jing Liao, Kieren Egan, Hanna M. Vesterinen, and Malcolm R. Macleod.

### Data analysis

Charis Wong, Jenna M. Gregory, Jing Liao, Kieren Egan, Hanna M. Vesterinen, and Malcolm R. Macleod.

### Methodology

Charis Wong, Jenna M. Gregory, Jing Liao, Kieren Egan, Hanna M. Vesterinen, and Malcolm R. Macleod.

### Software and programming

Jing Liao.

### Visualisation

Charis Wong.

### In vitro drug screening

Alessandra Cardinali, and Bhuvaneish T. Selvaraj.

### Expert panel

Jeremy Chataway, Robert Swingler, Peter Connick, Suvankar Pal, Siddharthan Chandran, and Malcolm R. Macleod.

### Writing - original draft

Charis Wong and Jenna M. Gregory.

### Writing - review & editing

Charis Wong, Jenna M. Gregory, Jing Liao, John Cafferkey, Arpan R. Mehta, Jeremy Chataway, Robert Swingler, Suvankar Pal, Siddharthan Chandran, and Malcolm R. Macleod.

All authors have read and approved the manuscript.

## DATA AVAILABILITY

Data are provided in supplementary material.

