## Supplemental data for "A Systematic Approach to Identify Neuroprotective Interventions for Motor Neuron Disease"

**A novel strategy to identify candidate drugs for clinical trial in Motor Neuron disease**

> **Note: This protocol was developed in 2012-13 and saved to a local machine on 25th May 2013. We now make it available, unamended save for this comment. The project has proceeded, with amendments to the protocol, and the reasons for these changes, will be described in an updated study protocol.**
>
> **Malcolm Macleod and Charis Wong, 16th September 2019**

**Aim**

To inform the selection of up to 4 putative neuroprotective oral interventions to be tested in an adaptive clinical trial of efficacy in patients with motor neuron disease

**Background**

Motor neuron disease (MND) is a rare neurological condition that leads to progressive muscle paralysis and death. While the condition is relatively uncommon (2 in 100 000 per year) the prognosis is extremely poor, with median survival from symptom onset of around three years. Current treatments have limited efficacy; the only drug known to increase life expectancy is riluzole, which only prolongs median survival by 3-4 months. While there has been a number of treatments developed which provide symptomatic benefit (e.g. non invasive ventilation) there is an urgent unmet medical need for interventions to improve life expectancy.

Systematic review and meta-analysis can be useful tools in the selection of candidate drugs for and the design of clinical trials. For example, systematic review and meta-analysis of animal and human data informed the design of a pragmatic multicentre €11m European trial of hypothermia for acute ischaemic stroke (www.eurohyp.org).

Similarly, the MS-SMART program set out to identify a short list of oral interventions which might be tested in a multi-arm phase IIb randomised double-blind clinical trial in progressive MS, based on efficacy both in cognate human conditions and in animal models of multiple sclerosis. After an initial systematic search to identify all interventions tested in MS and other diseases characterised by neurodegeneration a shortlist of 50 candidate interventions was drafted from 1374 original candidate interventions through the systematic application of predefined selection criteria.

All clinical studies describing the 50 shortlisted interventions acros the various conditions were assessed for reported efficacy, safety, study size and study quality and summary scores (graded from one (low scoring) through four (high scoring)) were generated for each of these dimensions. Additionally, we assessed reports of the efficacy of these shortlisted interventions in experimental autoimmune encephalomyelitis (EAE), the most common animal model of MS, using systematic review and meta-analysis. Finally, we presented all summary data at a round table discussion with a panel including clinical trialists, basic scientists, funders, patient groups and patient representatives. Through 2 cycles of exclusion we reduced the list to 20 and then to 7 drugs which might be used in clinical trial. These were ranked, and the trial steering committee then chose three drugs from these 7 to go forward. The trial, which has now been funded, will use change in MRI grey matter volume as a surrogate measure of clinical outcome.

Here we propose to use a methodology similar to that of MS-SMART to identify candidate drugs for clinical trial in Motor Neuron Disease.

**Approach**

We will use the comprehensive database of clinical trials of five neurological diseases (up to August 2011) collected in the context of MS-SMART, held by the CAMARADES collaboration and updated using the same strategy, to identify interventions which have been tested in MND at least once, or in at least three of Huntington's disease, Parkinson's disease, Alzheimer's disease or multiple sclerosis. Two independent reviewers (PC and SC) will use predefined criteria to identify potential candidate interventions.

We will then assess the efficacy, safety, size and quality of each publication testing an included intervention using the same methodology as for MS SMART. To complement the clinical data we will assess the efficacy of these interventions in animal models of MND. Finally we will produce summaries of the clinical (safety, efficacy, study size and study quality) and preclinical data for each intervention. This will then be presented for discussion at a consensus meeting to select from these, using the information presented along with their judgement and existing knowledge, intervention(s) which might be taken forward to clinical trial. As with MS-SMART, if there are interventions not included at this stage which panel members think should be considered these can be introduced and will, as far as is possible, be subject to the same rigorous standard of assessment.

**Methodology**

Our method to identify candidate oral interventions for a clinical trial on MND is to apply a transparent and reproducible 5-step process which is outlined in more detail below.

**1. Selection of publications to be included in the systematic review**

We have previously collated details of all clinical trials of oral interventions (up to August 2011) testing in multiple sclerosis (MS), Alzheimer's disease (AD), Parkison's disease (PD), Huntington's disease (HD) and motor neuron disease (MND) and amyotrophic lateral sclerosis (ALS) which is held on the CAMARADES Microsoft Access (2003) Data-Manager (www.camardes.info). Specifically we searched three online databases (PubMed, ISI Web of Knowledge, and Embase) as well as the UK clinical trials database (www.clinicaltrials.gov, accessed August 2011) for all clinical reports. We included case reports, uncontrolled case series, non randomised parallel group studies, crossover studies and randomised controlled trials and our search was not limited by date or language. We used the search terms: [motor neuron disease] OR [amyotrophic lateral sclerosis] OR [multiple sclerosis] OR [Alzheimer's disease] OR [Parkinson's disease] OR [Huntington's disease]. The search will be updates to take into account more recent publications.

Additionally we will screen the x database to identify any further relevant clinical trials including unpublished trials which they have identified.

Publications identified in the new search will be screened independently by two reviewers in Reference Manager against our pre-defined inclusion and exclusion criteria (Table 1). Data for publication ID, author, year of publication, intervention tested and disease will be extracted to the CAMARADES database for new publications meeting the inclusion and exclusion criteria. At each stage of the short-listing process, data will be extracted and analysed using this database.

| |
|---|
| **Inclusion criteria**<br>• Qualitative or quantitative data provided on either safety or efficacy for an orally delivered intervention, tested in any human clinical trial.<br>• Primary reporting of change in clinical status (including death, tracheostomy free survival, relapse frequency, disability progression, behavioural symptoms) or changes in biomarkers of clinical status (including magnetic resonance imaging (MRI), blood, cerebral spinal fluid (CSF), muscle strength). |
| **Exclusion criteria**<br>• Isolated reporting of non-pharamacological interventions such as acupuncture, aromatherapy, physiotherapy, or exercise.<br>• Articles reporting the use of interventions already licensed for clinical use for MND such as riluzole.<br>• Articles on levodopa treatment for Parkinson's disease.<br>• Studies reporting different modes of intervention delivery other than oral administration.<br>• Publications reporting secondary analysis of previously published clinical trial data.<br>• Protocols for potential clinical trials.<br>• Articles on relapsing-remitting MS.<br>• Combination treatments including where an oral and a non-oral intervention are administered. |

**Table 1.** *Eligibility criteria for publications included in systematic review*

**2. Stage 1 selection of interventions for retention in the review**

We will select interventions for further consideration if they have been tested in MND at least once, or tested in at least three of the four other diseases under consideration. As of May 2013 KE has identified our long list of interventions and is gathering relevant pdfs.

**3. Stage 2 selection of interventions for retention in the review.**

Next, two clinicians (SC & PC) will independently reviewed interventions retained from stage 1 to identify those that met pre-defined criteria of potential clinical applicability in MND and the absence of current or planned clinical trials in MND. The criteria are:

Inclusion criteria

1. The intervention is available orally
2. The drug would be available to investigators for clinical trial
3. A clinical trial using the drug would be feasible

Exclusion criteria

1. Efficacy, or lack of efficacy, is known with certainty in patients with MND.
2. Treatment is known to be associated with substantial harms that would outweigh reasonable predictions of potential benefits.
3. Known current or planned trial in MND.
4. Pragmatic consideration of what is known of the drug means that a clinical trial is unlikely to gain sufficient support from neurology clinical trialists, regulators or funders.

### 4. Stage 3: Systematic evaluation of efficacy, safety, amount of data and study quality for remaining interventions.

For retained interventions, publications will be assessed against predefined criteria by two of four investigators (SC, PC, SP, MM). Publications assessed in the context of MS-SMART will have these assessments carried forward..

Each report will be graded on a scale of one (worst) to four (best) for their reported evidence on safety and efficacy, study quality, and study size.

**Safety data** are scored as:

- "SUSARs (suspected unexpected serious adverse reactions) or unexpected mortality observed" or "not described" (1 point),
- "SAEs (serious adverse events) only" (2 points),
- "AEs (adverse events) only" (3 points)
- "no adverse effects reported" (4 points).

Each reported outcome will be given an **efficacy score** as:

- "not presented" or "definite worsening" (1 point)
- "neutral" (2 points)

- “non-significant improvement” (3 points)
- “significant improvement” (4 points).

The efficacy score for each outcome will be averaged for each publication to obtain an overall score.

Study quality will be assessed against a 21 point checklist comprising of a combination of criteria defined through a Delphi process,[18] GRADE,[19] and CAMARADES[20] methods.
A final publication-specific quality score of 1 to 4 will be allocated based on quartile membership (with quartiles defined by total quality scores across all publications):

- first quartile (1 point)
- second quartile (2 points)
- third quartile (3 points)
- fourth quartile (4 points).

**Study size** will be weighted as:

- “1-10 patients” (1 point)
- “11-100 patients” (2 points)
- “101-1000 patients” (3 points)
- “1001+ patients” (4 points).

Where a publication reports outcome in more than one experimental cohort (compared to a control group if present) an overall score for each publication will be generated by multiplying the average scores for efficacy, quality, safety, and study size for each experimental cohort. Where a number of outcomes are measured in the same experimental cohort we will extract data for the primary outcome if one was identified, and if not we will extract data for the most outcome showing greatest efficacy. Where there are different approaches to managing the risk of bias (eg some blinded and others unblinded) within an experimental cohort we will assess study quality according to the outcome which is taken forward under the efficacy score. We will also extract data for clinical trial type (*e.g.* RCT, un-controlled, cross-over etc.), clinical trial phase, mean age of the patients, sex of the patients, dose, duration of treatment, multi-centre or single-centre study, and the funding source.

Summaries will then be generated for each intervention including: number of patients, disease group(s) studied, design of studies, duration of studies, dose(s) administered, overall publication scores, and median scores for efficacy, safety, and quality.

"Heat maps" will be created by tabulating the number of publications awarded each of grades 1-4 for efficacy versus safety, efficacy versus quality and safety versus quality. An overall score for each intervention ("drug score") will be calculated as the product of the mean publication scores for efficacy, quality, safety and study size, and of the $\log_{10}$ of the number of publications identified plus 1.

### 5. Stage 4: Systematic evaluation of data from in vivo studies testing the efficacy of remaining interventions in animal models of MND

We will evaluate reports of the efficacy of remaining interventions in animal models of MND using Pubmed, Embase and ISI. For each experimental cohort we will extract data for time to treatment; the number of animals per group; mortality; functional outcome (e.g. neurobehavioural score) and structural outcome (e.g. motor neuron loss).
We will evaluate risk of bias based on a checklist previously described.[21] comprising the reporting of (1) random allocation to group; (2) blinded assessment of outcomes; (3) prior sample size calculation; (4) compliance with animal welfare regulations; (5) a statement of any potential conflicts of interest; and (6) inclusion and exclusion criteria.

### 6. Stage 5: Candidate drug selection committee review

A specially convened International MND Drug Selection meeting comprising expert representation from neuroscientists, neurologists, brain imaging, people with MND, trial methodologists and industry will consider the remaining drugs. If there are interventions not identified in the original search, or excluded at stages 1 or 2, but which panel members think should be considered, these can be introduced at this stage and will, as far as is possible, be subject to the same rigorous standard of assessment.

Structured discussions will occur over three rounds of detailed scrutiny. Remaining interventions will be eliminated through an iterative process based on review of information from Stage 3 and Stage 4 and with consideration of biological plausibility, pharmacological criteria such as CNS penetration and presumed mechanistic class of action. This process will be concluded by identification of a final group, to be ranked and categorised according to class of action and mechanistic plausibility for effects on pivotal neurodegenerative pathways.
